# Exploring the common genetic architecture of autism spectrum disorder using a novel multi-polygenic risk score approach

**DOI:** 10.1101/2023.05.23.23290405

**Authors:** Zoe Schmilovich, Vincent-Raphaël Bourque, Guillaume Huguet, Qin He, Jay P. Ross, Martineau Jean-Louis, Zohra Saci, Boris Chaumette, Patrick A. Dion, Sébastien Jacquemont, Guy A. Rouleau

## Abstract

Compared to disorders of similar heritability and contribution of common variants, few genome-wide significant loci have been implicated in autism spectrum disorder (ASD). This undermines the use of polygenic risk scores (PRSs) to investigate the common genetic architecture of ASD. Deconstructing PRS-ASD into its related traits via “developmental deconstruction” could reveal the underlying genetic liabilities of ASD. Using the data of >24k individuals with ASD and >28k of their unaffected family members from the SSC, SPARK, and MSSNG cohorts, we computed the PRSs for ASD and 11 genetically-related traits. We applied an unsupervised learning approach to the ASD-related PRSs to derive “multi-PRSs” that captured their variability in orthogonal dimensions. We found that multi-PRSs captured a similar proportion of genetic risk for ASD in cases versus intrafamilial controls (OR_multi-PRS_=1.10, R^2^=0.501%), compared to PRS-ASD itself (OR_PRS-ASD_=1.16, R^2^=0.619%). While multi-PRS dimensions conferred risk for ASD, they had “mirroring” effects on developmental phenotypes among cases with ASD. We posit that this phenomenon may partially account for the paucity of genome-wide significant loci and the clinical heterogeneity of ASD. This approach can serve as a proxy for PRS-ASD in cases where non-overlapping and well-powered GWAS summary statistics are difficult to obtain, or accounting for heterogeneity in a single dimension is preferable. This approach may also capture the overall liability for a condition (*i.e*.: genetic “P-factor”). Altogether, we present a novel approach to studying the role of inherited, additive, and non-specific genetic risk factors in ASD.

## INTRODUCTION

Autism spectrum disorder (ASD) has a high heritability ranging from 0.65 to 0.91^1–3^. Rare *de novo* copy-number variants (CNVs) and single-nucleotide variants (SNVs) in highly constrained genes of large effect sizes have been implicated in ASD, yet they collectively explain <5% of the overall liability for the disorder^4–6^. In contrast, it is estimated that >50% of the liability for ASD resides in common variation^4–9^. Despite its large contribution to genetic risk, the common variant genetic architecture of ASD remains elusive.

Genome-wide association studies (GWASs) have been instrumental in identifying common risk loci in disorders^10^. Despite the substantial increase in the sample size of ASD GWASs over the last decade^11–16^, only five genome-wide significant loci have been implicated in ASD to date^17^. The number of risk loci identified in ASD is markedly lower than in other psychopathologies. For instance, schizophrenia has a similar narrow sense heritability (81%)^18^ and an estimated contribution of common genetic variation that is comparable to ASD^19^. However, >100 genome-wide significant loci have been implicated in schizophrenia. One potential explanation for this discrepancy is the difference in study sample sizes between the two disorders: the schizophrenia GWAS includes more cases than the ASD GWAS (*i.e*.: >36k cases in schizophrenia versus 18k in ASD). Some projections on the genetic architecture of these disorders suggest that ∼70k cases could suffice to yield 100 genome-wide significant loci in ASD^20^. However, this would still represent a steep improvement curve, and recent analyses do not appear to follow such projections^21^. This questions whether other factors besides solely sample sizes could contribute to the lower number of implicated loci in ASD.

We hypothesize that the complex and heterogeneous genetic architecture of ASD may partially explain the paucity of genome-wide significant hits. The scarcity of ASD GWAS hits may, in turn, limit the accuracy of polygenic risk scores (PRSs), which rely on summary statistics of variant associations to generate individual-level genetic susceptibility scores for a trait^22^.

The phenotypic overlap^23, 24^ and genetic correlation^17, 25, 26^ between ASD and diverse developmental phenotypes have been well-documented. In fact, it has been suggested that ASD may arise as a result of increased inherited genetic susceptibility for various developmental phenotypes^27^. As such, we sought to study the genetic architecture of ASD via “developmental deconstruction”, *i.e.*: deconstructing the unitary ASD syndrome liability into its contributory developmental phenotypes, both ASD-specific and non-specific factors^28^. Mous and colleagues previously showed that background susceptibilities for ADHD and for motor coordination that are inherited, associated, but non-specific to ASD (referred to as “BASINS”), may contribute to the additive genetic liability for ASD in the same way that ASD-specific risk factors could contribute to a diagnosis for ASD^27^. More recently, Warrier and colleagues showed that common variants associated with ADHD and educational attainment (*i.e.*: non-ASD-specific risk factors) contribute to several core features of ASD^29^. However, these studies have focused on univariate associations between the polygenic risk of one trait (ASD) and its core symptoms. Previous studies did not account for the fact that many genetic variants are represented – at different degrees of association or weighting – in more than one of these PRSs. These studies also did not account for the effect of these genetic liabilities on developmental phenotypes outside of the core symptoms of ASD. Accounting for the effect of multiple ASD-related PRSs on ASD risk and related developmental phenotypes in a single model is a knowledge gap that remains unexplored.

We posit that modelling ASD and its associated features as a function of multiple ASD-related PRSs via developmental deconstruction could: 1–serve as a proxy for PRS-ASD, and; 2–highlight the heterogeneity underlying the common genetic architecture of ASD.

To do this, we analyzed a sample of 28,307 cases with ASD and 50,953 of their unaffected relatives across three ASD cohorts. First, we computed the PRSs for ASD and 11 of its genetically correlated traits, as reported in the Grove *et al.* ASD GWAS^17^. Then, we applied an unsupervised learning algorithm (principal component analysis, PCA) to construct PRSs that captured the variation of the 11 ASD-related traits across orthogonal principal components (PCs) that we refer to as “multi-PRS” dimensions.

This study provides support for the use of multi-PRS dimensions (constructed from ASD-related traits) to capture the additive, inherited, and non-specific genetic liability for ASD. First, we showed that the multi-PRS dimensions can capture a similar proportion of the inherited genetic liability of ASD risk compared to PRS-ASD itself (0.501% versus 0.619%). Second, we modelled the effect of PRS-ASD and multi-PRS liability on 46 developmental phenotypes among cases with ASD. Our results reveal that the multi-PRS dimensions can capture unique phenotypic differences among the cases with ASD that PRS-ASD cannot. Interestingly, while multi-PRS dimensions increased the risk for ASD, they had “mirroring” effects on core ASD symptoms, developmental features, and the risk for co-occurring disorders. These findings provide support for the clinical and genetic heterogeneity of ASD, which in turn, may partially explain the paucity of reproducible ASD GWAS hits.

## MATERIALS AND METHODS

### Cohorts

In total, the genetic data of 28,307 individuals with a diagnosis of ASD and 50,953 of their unaffected siblings and parents were included in this study using the available data across three family-based ASD cohorts: the Simons Simplex Collection (SSC)^30^, Simons Foundation Powering Autism Research for Knowledge (SPARK)^31^, and MSSNG^32^ (Table S1). We excluded 546 parents and siblings with a diagnosis of ASD from the study.

### Genetic data quality control

We performed quality control of the genetic data for each platform (SNP genotyping: Illumina 1Mv1, 1Mv3, Omni2.5 for SSC; Illumina Infinium Global Screening Array-24 for SPARK, and Whole-Genome Sequencing (WGS): Complete Genomics and Illumina HiSeq (2000 and X Ten) for MSSNG) separately. Standard quality control filtering criteria were applied to the genetic data^33^. We excluded individuals with genotyping rate <95%, excessive heterozygosity (± 3 standard deviations from the mean), sample missingness >0.02, mismatched in reported and genetic sex, and families with Mendelian errors >5%. We removed SNPs with a call rate <98%, a minor allele frequency (MAF) <1%, deviated from Hardy-Weinberg Equilibrium (P <1×10^-6^), and >10% Mendel error rate.

We used the *mds* parameter from the KING software^34^ to infer the population substructure of the samples in the study. To avoid confounders due to ancestry, we restricted our analyses to only individuals with a >90% probability of inferred-European ancestry. Imputation of SNP genotypes was performed using the 1000 Genomes Project, phase 3 (1KGP3) reference panel^35^ through the Sanger Imputation Server (https://www.sanger.ac.uk/tool/sanger-imputation-service/). The VCFs of the imputed SNP and WGS genotypes were subsequently merged using the *merge* command from the “bcftools” program^36^, such that only the loci that were present across all technologies were retained^36^. The merged imputed files were then converted to PLINK files, and were subsequently filtered to remove SNPs with a poor imputation quality (≥0.3); more than 2 alleles (multiallelic variants); MAF <5%; call rate <98%, and deviated from Hardy-Weinberg Equilibrium (P < 5×10^-7^). Finally, we computed the top 10 ancestry principal components (PCs) for the final European samples using the “mds” parameter from the KING software^34^.

Following sample and variant-level quality control, 24,549 individuals with ASD and 28,898 unaffected family members were retained in the study. A summary and description of the final samples included in the study are detailed in Table S2.

### Polygenic risk score (PRS) calculation

PRSs were constructed using the GWAS summary statistics of ASD and 11 other traits with a reported significant genetic correlation with ASD^17^ (Table 1). To avoid sample overlap, custom summary statistics for the ASD GWAS summary statistics were generated to exclude the SSC cohort (obtained through the PGC application for secondary analysis proposal – https://pgc.unc.edu/for-researchers/data-access-committee/data-access-information/).

**Table 1.**
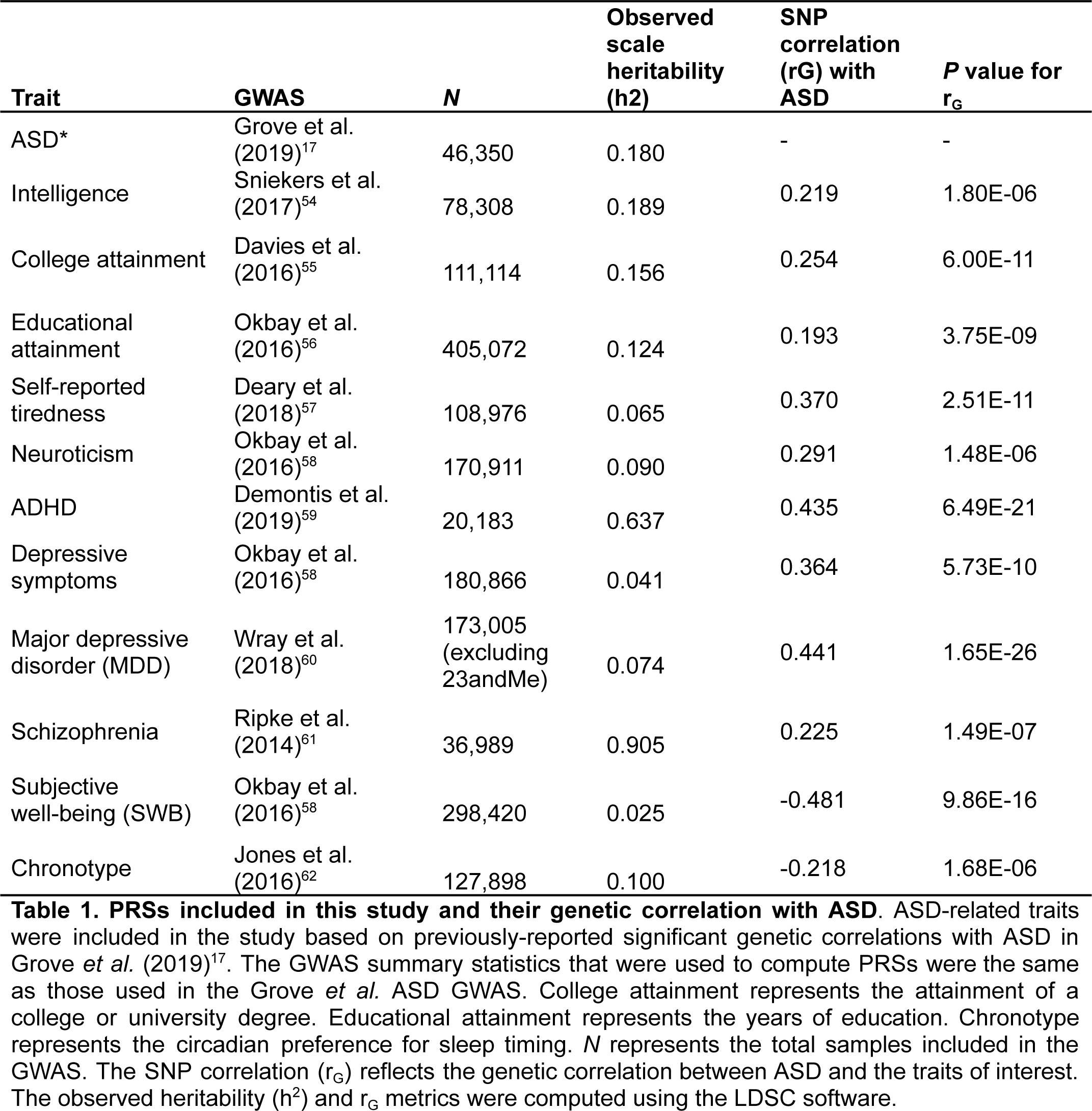
PRSs included in this study and their genetic correlation with ASD. ASD-related traits were included in the study based on previously-reported significant genetic correlations with ASD in Grove *et al.* (2019)^17^. The GWAS summary statistics that were used to compute PRSs were the same as those used in the Grove *et al.* ASD GWAS. College attainment represents the attainment of a college or university degree. Educational attainment represents the years of education. Chronotype represents the circadian preference for sleep timing. *N* represents the total samples included in the GWAS. The SNP correlation (r_G_) reflects the genetic correlation between ASD and the traits of interest. The observed heritability (h^2^) and r_G_ metrics were computed using the LDSC software.

We used PRS-CS^37^ to infer the posterior effect sizes of SNPs using GWAS summary statistics and the 1KGP3 European linkage disequilibrium (LD) reference panel^35^. Individual-level polygenic risk scores were then computed from the PRS-CS output summary statistics using the PLINK^38^ “score” and “no-mean-imputation” parameters.

To account for subtle differences in population structure among the samples with inferred European ancestry, the PRS of each trait was modelled as a function of the top 10 ancestry principal components (PCs) in a linear regression (*lm* function within the R *stats* package): **PR*S*_trait_ ∼ **PC**_1_ + **PC**_2_ + **PC**_3_ + … + **PC**_10_. Then, the residuals from each regression model were extracted to represent PRS values that accounted for the underlying effects of ancestry among the samples. Finally, the PRSs for each trait were transformed into z-scores.

### Correlation between ASD and related psychopathologies

The genetic correlation (r_G_) between ASD and its related psychopathologies were computed using the command-line tool LD SCore (LDSC, v1.0.1)^39, 40^ based on the included GWAS summary statistics (Table 1). The correlation between the PRSs (Figure 2) was computed using the *cor* functions from the “stats” base R package.

### Reducing PRSs of ASD-related traits into representative principal components

PCA is a dimensionality reduction technique used to compress multidimensional data into representative principal components (PCs) while retaining the most amount of information. We employed this technique to reduce the PRSs of the 11 ASD-correlated traits (Table 1) into variables that captured the variability of all the traits into single dimensions (PCs). We refer to these “reduced” PC variables – representing an individual’s genetic propensity for all 11 ASD-related traits – as “multi-PRS” dimensions. We used this approach to model the effect of all ASD-related traits in a single regression. Given that the PCs from the PCA are orthogonal, we can include all multi-PRS variables as predictors without violating the regression assumption of the absence of multicollinearity.

To do this, we used the *PCA* function to perform the PCA, and the *get_pca* function to extract the output from the “factoextra” R package^41^. The results from this analysis are detailed in Figure 3. As a sensitivity analysis, we also ran the PCA within six different subgroups (cases with ASD, intrafamilial controls, and the three separate cohorts) (Figures S4, S5, S6). The sensitivity analyses confirm that the PC loadings discussed in the main analyses, whereby we group all samples together, are not driven by any one of these subgroups.

### Statistical analyses

#### Effect of PRS dimensions on ASD risk in cases versus their unaffected family members

The effect of the PRSs on ASD risk in cases with ASD versus their unaffected family members was modelled using a generalized linear mixed-effects (GLME) model using the *glmer* function from the “lme4*”* R package (see Figure 1 for the analysis workflow). This model accounts for the effects of relatedness among ASD individuals and their intrafamilial controls by including the family identifier as a random effect. We ran 13 separate GLME models with ASD diagnosis as the outcome, with the following predictors:

**Figure 1.**
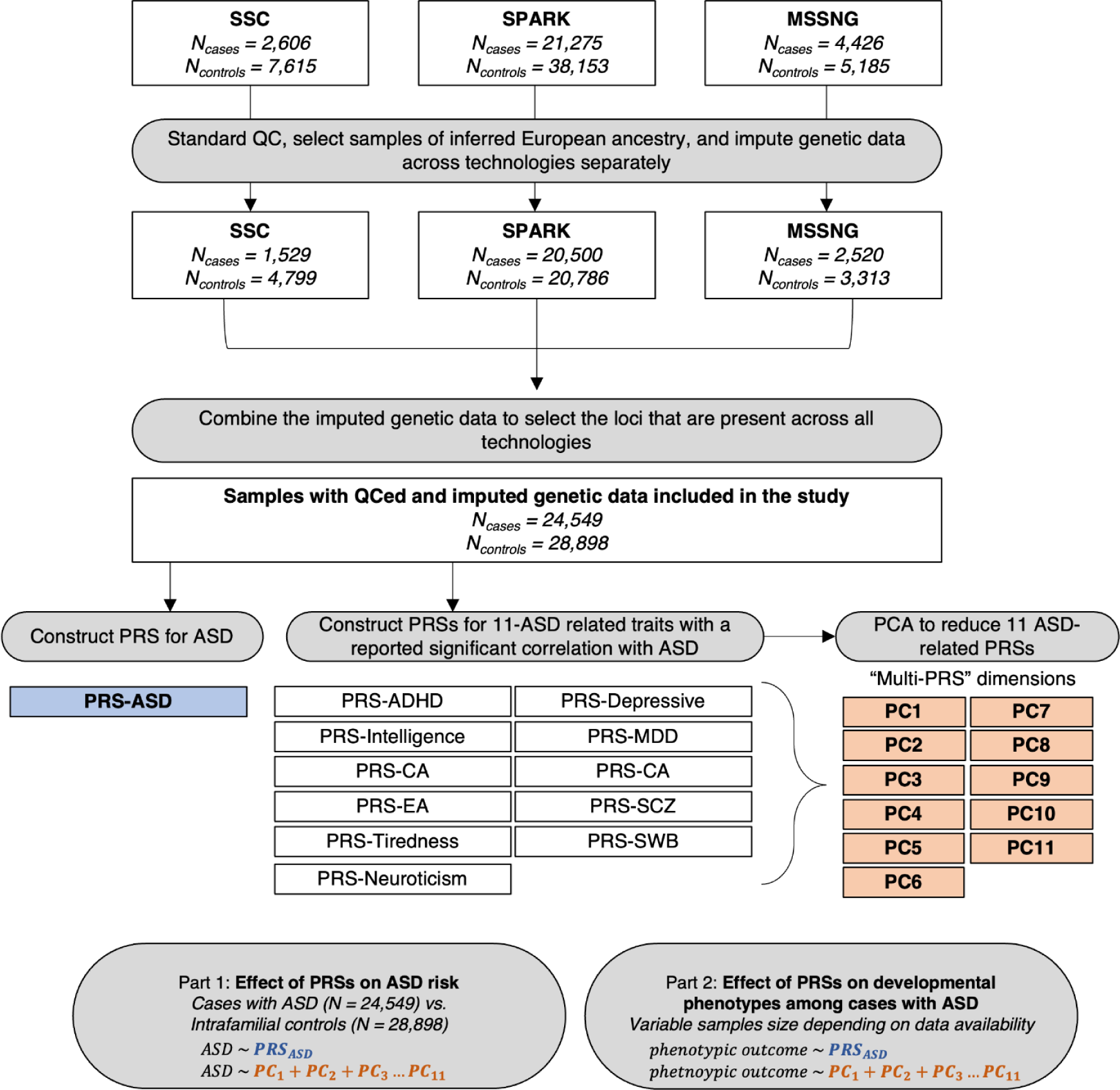
Analysis flowchart. The genetic data of three family-based ASD cohorts were included in the study. Following standard genetic and sample-level QC, 24,549 cases with ASD and 28,898 of their unaffected family members (intrafamilial controls) were included in the study. Twelve separate polygenic risk scores (PRSs) were constructed using the GWAS summary statistics of ASD and the 11 traits with a reported significant genetic correlation with ASD. PCA was applied to the 11 ASD-related traits to create “multi-PRS” dimensions (PCs). The first part of the study assessed the effect of the PRS dimensions on ASD risk in cases with ASD versus their unaffected family members. The second part of the study assesses the effect of the PRS dimensions on various developmental phenotypes among cases with ASD.

Model 1 (Naïve model using PRS-ASD as the predictor):

**ASD* ∼ *PRS*_*ASD*_*

Models 2–12 (PRSs of ASD-related traits as predictors):

**ASD* ∼ *PRS*_*ADHD*_*

**ASD* ∼ *PRS*_*MDD*_*

**ASD* ∼ *PRS*_*SWB*_*

**ASD* ∼ *PRS*_*tiredness*_*

**ASD* ∼ *PRS*_*SCZ*_*

*^*ASD* ∼ *PRS*^_*chronotype*_*

**ASD* ∼ *PRS*_*IQ*_*

**ASD* ∼ *PRS*_*EA*_*

**ASD* ∼ *PRS*_*CA*_*

**ASD* ∼ *PRS*_*neuroticism*_*

**ASD* ∼ *PRS*_*depressive*_*

Model 13 (multi-PRS dimensions that reflect the reduced PRSs of the 11 ASD-related traits): **ASD** ∼ **PC**1 + **PC**2 + **PC**3 + **PC**4 +**PC**5 + **PC**6 + **PC**7 + **PC**8 + **PC**9 + **PC**10 + **PC**11

For all the models above, sex was included as a covariate and familial relationship was included as a random effect variable.

The goodness-of-fit of each model was evaluated according to its R^2^ (detailed model performance metrics in Table S6). We computed the conditional R^2^ of each model with the *r.squaredGLMM* function from the “MuMIn” R package^42^. The conditional R^2^ represents the variance explained by the entire model, including both fixed and random effects. In this study, we were interested in the proportion of ASD risk that was captured by the PRS predictors in each model. To do this, we first ran a GLME regression that included only the covariates as predictors. Then, we subtracted the R^2^ of the covariate model from the conditional R^2^ of each PRS regression model (“*r.squared.adj”* in Table S6). For ease of interpretability, the adjusted R^2^ was multiplied by 100% to indicate the percentage of ASD variability that each model captured.

### Effect of PRS dimensions on developmental phenotypes in cases with ASD

To assess the effect of PRS-ASD and the multi-PRS dimensions on developmental phenotypes in cases with ASD, we used either a linear (*lm* function) or a logistic (*glm* function) regression model depending on the continuous or binary phenotypic outcome, respectively. The phenotypes were included if there were data available for ≥5% of the samples with ASD. Given the limited data availability, the number of ASD samples with phenotypic data ranged from 1484 (6.04%) to 21,857 (89.03%). Overall, 46 traits (18 continuous and 28 binary) were included in the analyses (Table S3). The continuous outcomes were standardized (Z-score) by the mean within each of the three cohorts. These developmental phenotypes were grouped into nine categories: core ASD features (3); cognitive ability (3); adaptive functioning (3); developmental features (11); co-occurring disorders (17); language ability (4); family history (2); neurological disorder (1), and; health outcome (1). In total, we ran 92 regression models: two models for each developmental phenotype, using either PRS-ASD or all multi-PRS dimensions as the predictors. All models were adjusted for sex and age. The detailed performance metrics of each model are detailed in Table S8.

All *P* values were adjusted by the Benjamini–Hochberg false-discovery rate (FDR) correction for multiple comparisons using the *p.adjust* function from the base R package.

## RESULTS

### Genetic and PRS correlation between ASD and ASD-related traits

The results from the genetic and PRS correlation between ASD and its related traits are detailed in Figure 2. We included all 11 traits that had a reported significant genetic correlation with ASD from the latest ASD GWAS^17^. Ten traits (ADHD, MDD, depressive symptoms, tiredness, neuroticism, college attainment, schizophrenia, intelligence, and educational attainment) had a significant genetic (r_G_) and PRS correlation with ASD (P_FDR_ < 0.05). Two traits (chronotype and subjective well-being) had a significant negative genetic (r_G_) and PRS correlation with ASD. The genetic and PRS correlations in our study were concordant with those reported in Grove *et al.* This is expected, given that PRSs we computed were derived from the GWAS summary statistics used to compute the r_G_ in the ASD GWAS17. This finding highlights the shared genetic heterogeneity among all traits included in this study. Indeed, every trait that has a significant genetic correlation with ASD is also correlated with the PRS of another ASD-related trait. The high correlation between ASD and the ASD-related traits supports the use of a dimensionality reduction approach to flatten the variability of the PRSs into orthogonal variables.

**Figure 2.**
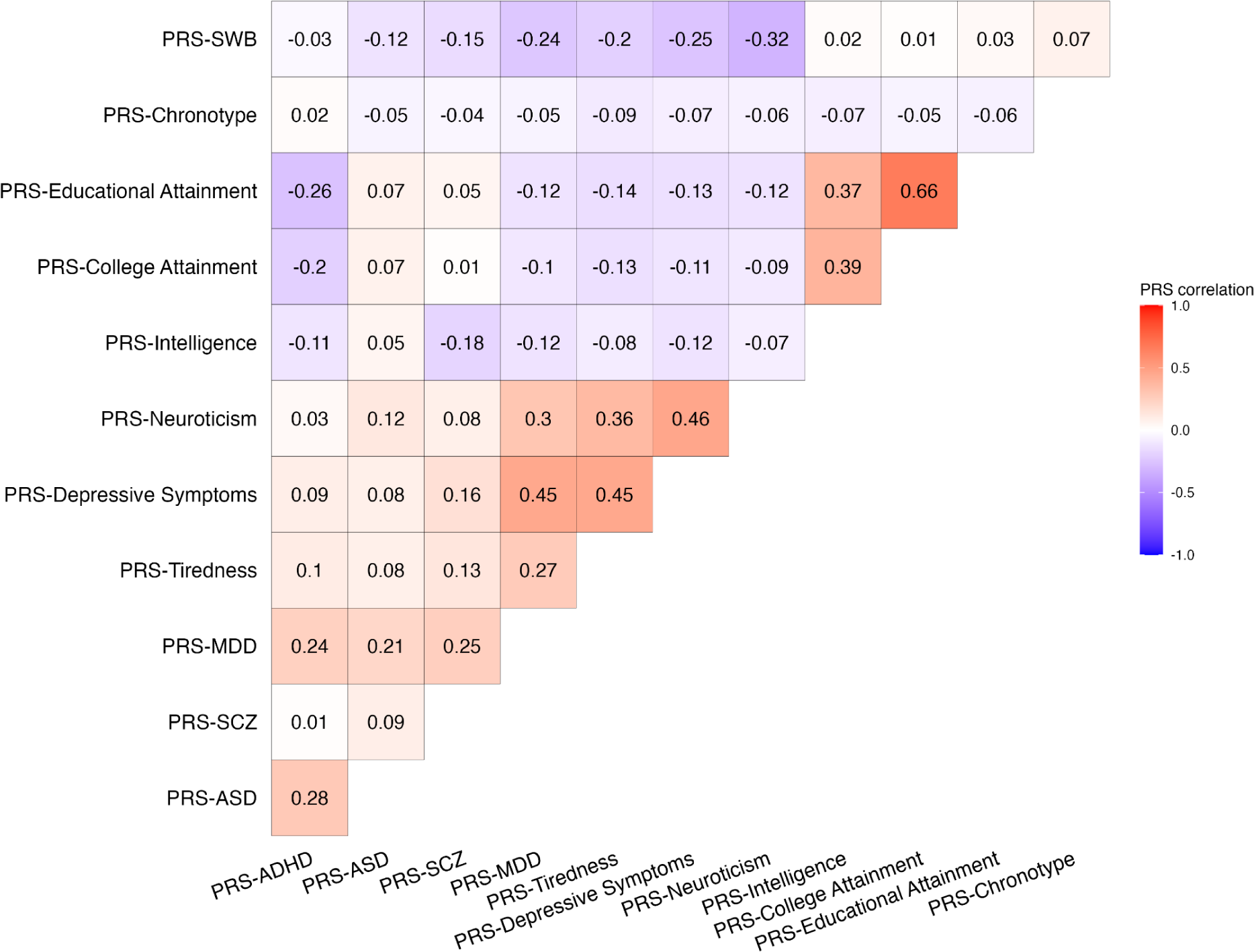
Genetic relationship between ASD and its related traits. The correlation between PRS-ASD and the PRS of its 11 related traits.

### PRSs for distinct ASD-related traits can be reduced to representative “multi-PRS” dimensions

Given the correlation between PRS-ASD and its related traits, we assessed whether multi-PRS dimensions, which encompassed the variability of polygenic risk across 11 ASD-related traits, could be used as a proxy to capture the genetic risk for ASD. We used PCA to capture the variability of PRS across all (n=11) ASD-related traits into representative “multi-PRS” dimensions (principal components, PCs).

The proportion of PRS variability captured by each PC ranged from 24.7% (PC1) to 3.1% (PC11) (Figure 3). The proportion of polygenic risk for each ASD-related trait that each multi-PRS dimension captures is detailed in Figure 4a. The correlation between PRS-ASD and each of the multi-PRS dimensions is detailed in Table S5.

**Figure 3.**
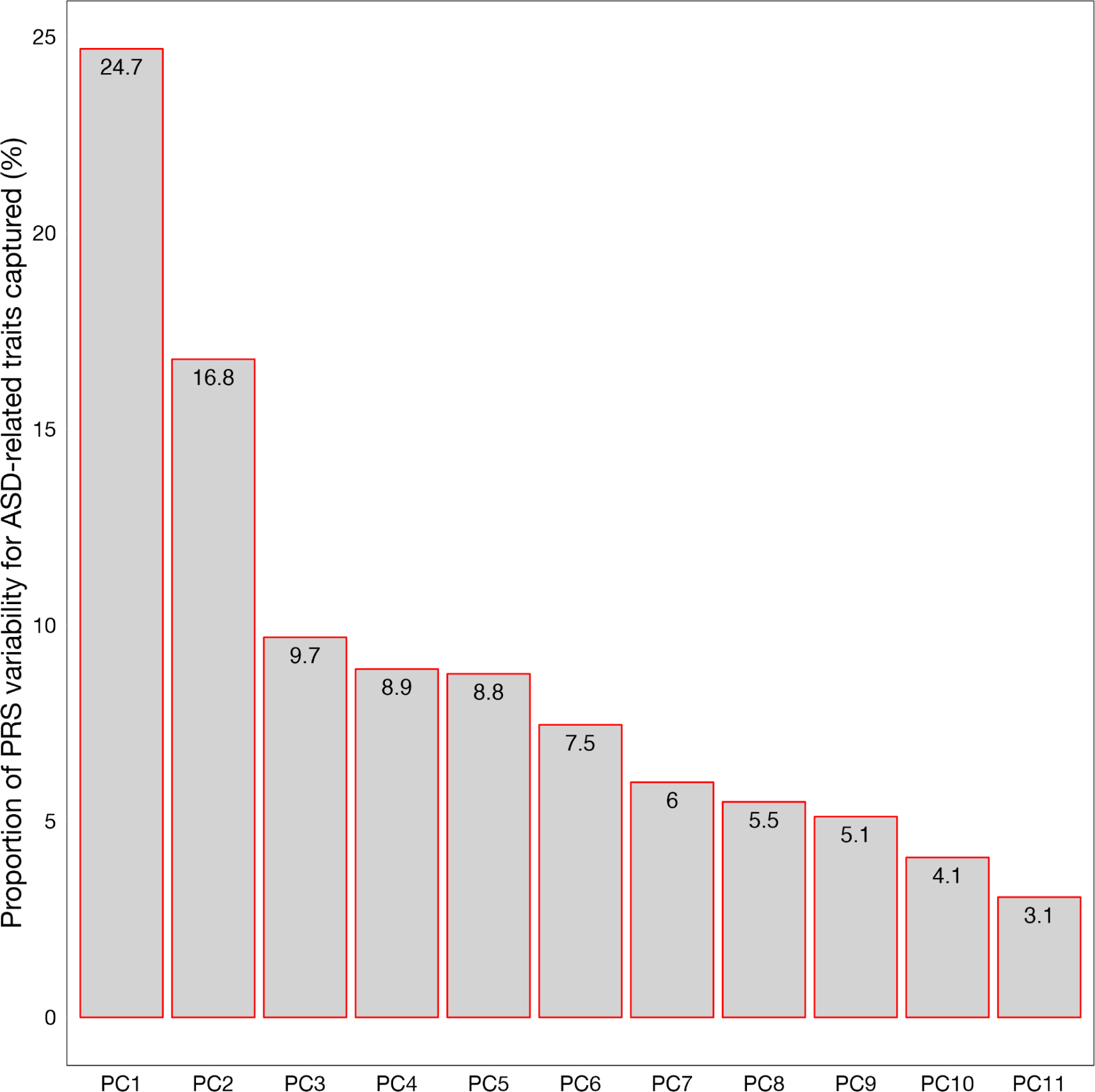
The proportion of PRS variability captured by each multi-PRS dimension. PCA was applied to the PRSs of the 11 ASD-related traits. Each resulting principal component (PC) represents the proportion (%) of PRS variability captured across all traits.

**Figure 4.**
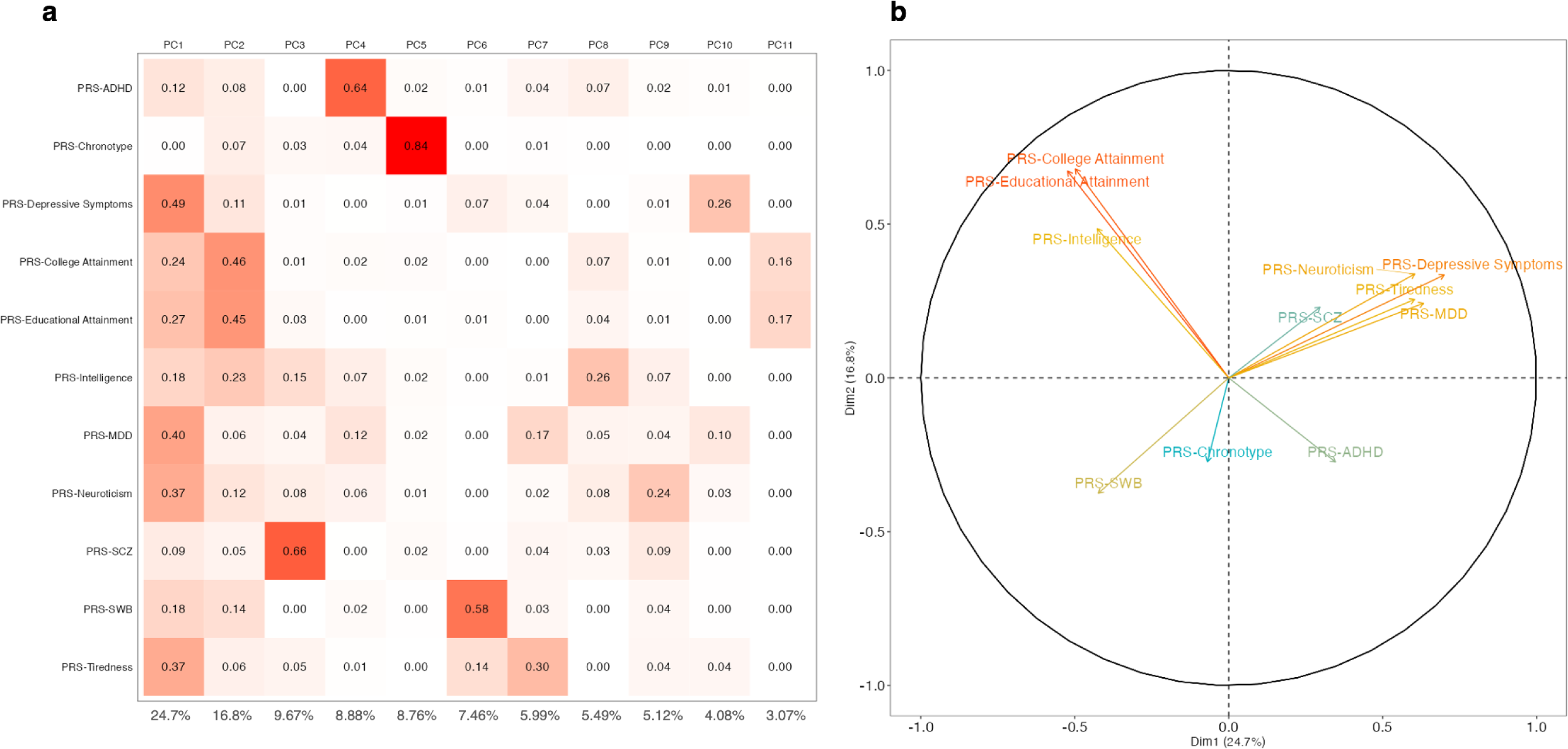
Reducing the 11 ASD-related PRSs into representative principal components (PCs), herein referred to as “multi-PRS” dimensions. **a**) The proportion of PRS variance that is captured in each multi-PRS PC dimension. The percentage of explained variance that each multi-PRS PC dimension captures is shown below each column. **b**) A loading plot representing the influence of each trait on PC1 (Dim.1) and PC2 (Dim.2). The traits that cluster together (similar contributions to PC1/PC2) have relatively similar PRSs.

Multi-PRS PC1 mostly captures the polygenic variability of negative symptom outcomes (i.e.: depressive symptoms, MDD, neuroticism, tiredness), whereas multi-PRS PC2 mostly captures the polygenic variability of higher cognitive ability (i.e.: college and educational attainment, intelligence). The similarities between the PRSs are represented in Figure 4b, whereby the traits that cluster together have similar contributions to PC1 and PC2.

Overall, these findings suggest that despite their high genetic and PRS overlap (Figure 2), there is a distinct pattern of genetic relatedness between the traits (Figure 4b). Moreover, these findings show that ASD-related PRSs can be transformed into representative – and orthogonal – multi-PRS dimensions that capture a substantial proportion of polygenic variability (Figure 3, Figure 4a). This, in turn, provides support for the potential use of multi-PRS dimensions as a proxy for PRS-ASD to capture the genetic risk for ASD.

### Multi-PRS dimensions capture a similar proportion of ASD risk, compared to PRS-ASD itself

We then modelled the effect of each PRS and the effect of the multi-PRS dimensions (*i.e.,* the PCs representing the variability across polygenic risk for the 11 ASD-related traits) on ASD risk in cases with ASD versus their unaffected family members (Figure 5a). An increase in polygenic risk for ASD, ADHD, MDD, tiredness, neuroticism, depressive symptoms, and schizophrenia significantly increased the risk for ASD diagnosis in probands compared to their unaffected family members. Individuals with ASD had a significant decrease in polygenic risk for subjective well-being and educational attainment in comparison to their unaffected family members. As expected, PRS-ASD had the highest effect on ASD risk (adjusted *P* value = 4.45E-52; OR = 1.16 [1.14, 1.18]).

**Figure 5.**
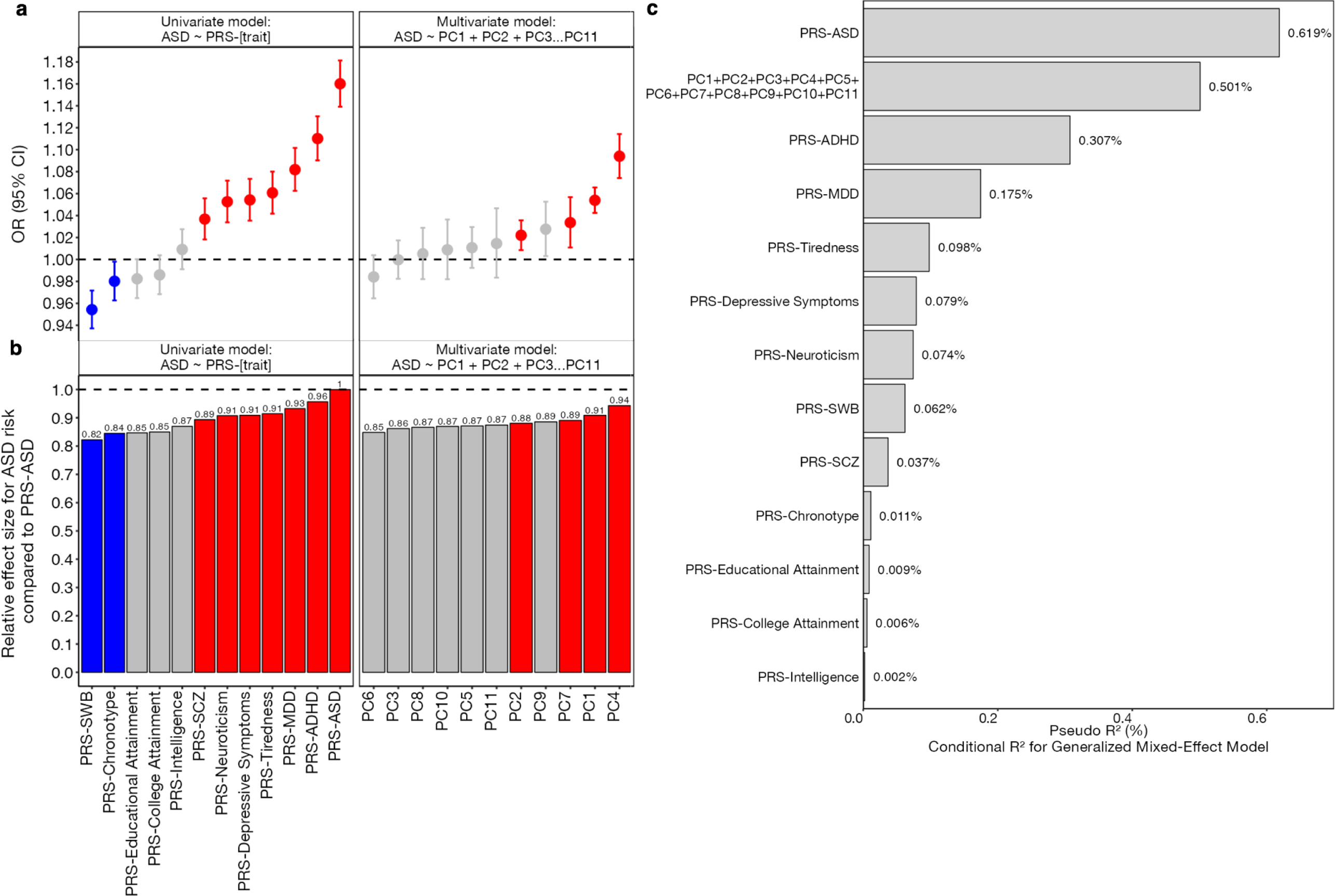
Assessing the effect of different PRS modalities on ASD risk. **a**) The effect of PRS-ASD, the PRS of ASD-related traits, and multi-PRS dimensions on ASD risk in cases versus their unaffected family members. The effect of PRS-ASD and PRS of ASD-related traits were modelled separately in univariate generalized linear mixed-effects models. Given that the multi-PRS dimensions were orthogonal, they were included as additive predictors in a single multivariate generalized linear mixed-effects model. Relatedness was included as a random effect in all models. Seven PRSs significantly increased the risk for ASD, and two PRSs significantly decreased the risk for ASD. Four of the multi-PRS dimensions significantly increased the risk for ASD. **b**) The relative odds ratio (OR) of each PRS and multi-PRS dimension on ASD risk, in comparison to PRS-ASD. The relative effect is represented as the proportion of OR relative to the effect of PRS-ASD (OR = 1.16). Red and blue points denote a significant (adjusted *P* value <0.05) positive or negative effect on ASD risk, respectively. All *P* values were adjusted for FDR correction for multiple comparisons. **c**) The proportion of ASD risk (conditional R^2^) that each regression model captures. The R^2^ represents the percentage of ASD risk that is captured by the PRS dimensions themselves. The multi-PRS model captures a similar proportion of ASD risk compared to the naive PRS-ASD model itself (0.501% versus 0.619%). The effects due to covariates and familial random effects were subtracted from this estimate. This suggests that the multi-PRS dimensions can be used as a proxy for PRS-ASD itself.

Four of the multi-PRS dimensions (PC1, PC2, PC4, and PC7) significantly increased the risk for ASD (P_FDR_ < 0.05) (Figure 5a). PC4 significantly increased the risk of ASD by 1.10 (adjusted *P* value = 2.81E-21; 95% CI of OR = [1.07, 1.11]) and captured 8.9% of variability across all PRSs (Figure 3) – including mostly the polygenic risk for ADHD (64%), MDD (12%), intelligence (7%), and neuroticism (6%) (Figure 4a). PC1 nominally increased the risk of ASD by 1.05 (adjusted *P* value = 2.44E-20; 95% CI of OR = [1.04, 1.07]) and captured 24.7% of the variability across all the PRSs (Figure 3), which mostly represented the variability of polygenic risk for depressive symptoms (49%), major depressive disorder (40%), neuroticism (37%), tiredness (36%), educational attainment (27%), and college attainment (25%) (Figure 4a). PC2 also nominally increased the risk for ASD by 1.02 (adjusted *P* value = 3.59E-03; 95% CI of OR = [1.01, 1.04]) and captured 16.8% of the variability across all PRSs (Figure 3), which mostly encompassed the variation of PRS for college attainment (46%), educational attainment (45%), and intelligence (23%) (Figure 4a).

In comparison, PRS-ASD increased the risk of ASD by 1.16 (adjusted *P* value = 9.60E-58; 95% CI of OR [1.14, 1.18]). The relative effect size of the significant multi-PRS dimensions, compared to PRS-ASD, ranged from 0.88 to 0.94 (Figure 5b).

We then compared the performance (R^2^) of the models according to the proportion of genetic risk for ASD that they captured (Figure 4C). The univariate regression model with PRS-ASD as the predictor (naive model) captured the greatest proportion of ASD risk (Conditional R^2^ = 0.619%). The multivariate regression model, which included all multi-PRS dimensions as predictors, captured a similar proportion of ASD risk (Conditional R^2^ = 0.501%) compared to the naive model. Importantly, given that each multi-PRS dimension is orthogonal, including them all as predictors would not violate any assumption of independence. In contrast, the univariate regression models that used each individual ASD-related PRS as separate predictors captured markedly lower proportions of ASD risk (ranging from 0.002% to 0.37%). The results from the multivariate regression that included PRS-ASD and all ASD-related PRSs as predictors is detailed in Table S7.

### Multi-PRS dimensions capture developmental variability among cases with ASD

We then assessed the effect of PRS-ASD and all multi-PRS dimensions on various developmental phenotypes among the samples with ASD (Figure 6).

**Figure 6.**
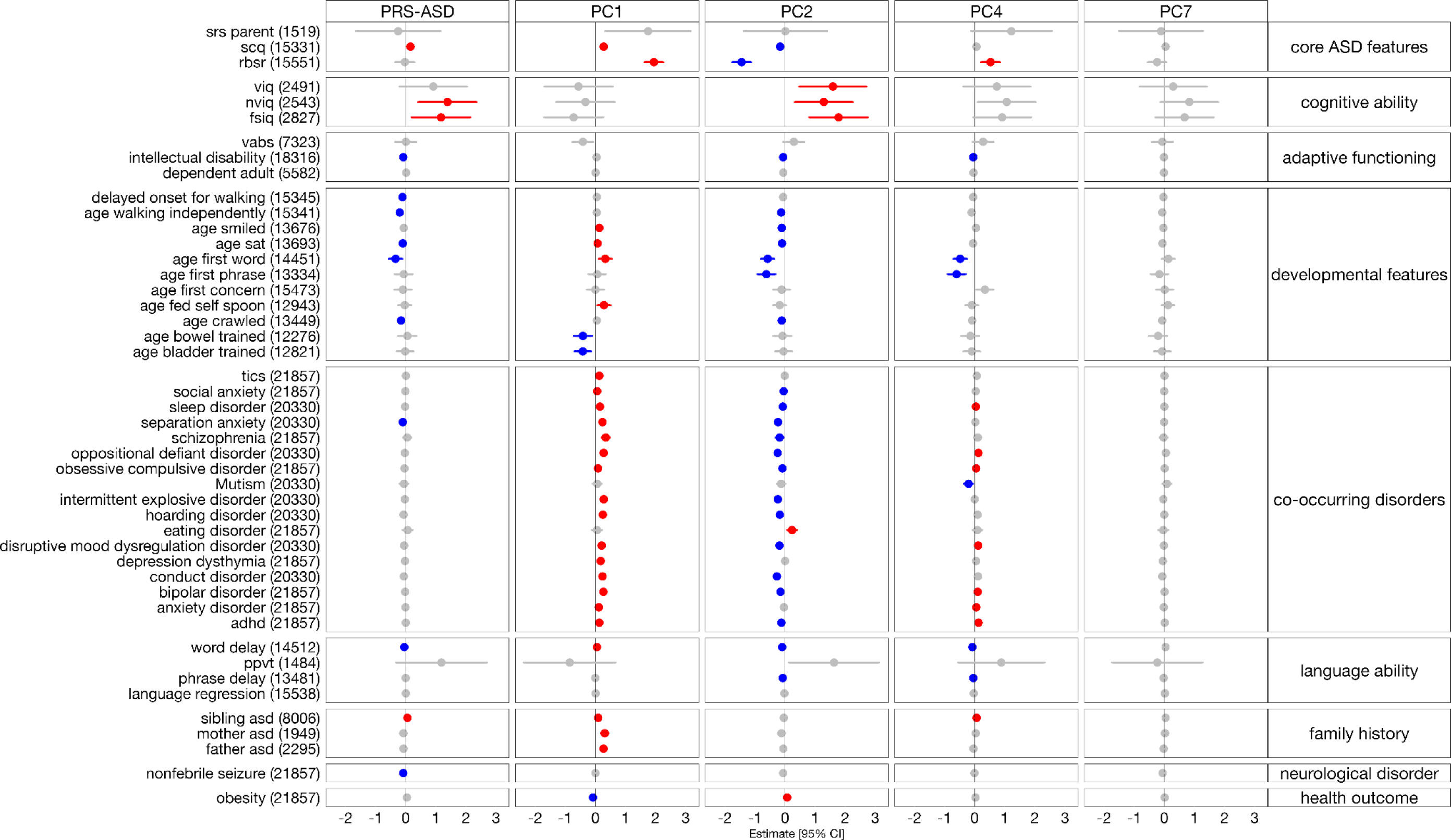
The effect of PRS-ASD and the multi-PRS dimensions on developmental phenotypes in cases with ASD. The results of 92 regressions are shown (46 outcomes with either PRS-ASD or all 11 multi-PRS dimensions as the predictors). While all 11 multi-PRS dimensions were included in the model, only those that had a significant effect on ASD risk (Figure 4a) are shown. Red and blue dots represent a significant positive and negative effect of the PRS predictor variable on the developmental phenotype, respectively. All *P* values were adjusted for FDR correction. The brackets denote the number of cases with ASD that were included in the model. The detailed statistical output of each model is detailed in Table S8. All models were adjusted for age and sex. Three and 13 comparisons were considered for the FDR correction of the PRS-ASD and multi-PRS models, respectively. SRS: Social Responsiveness Scale, SCQ: Social Communication Questionnaire, RBSR: Repetitive Behaviour Scale, VIQ: Verbal IQ, NVIQ: Nonverbal IQ, FSIQ: Full-scale IQ, VABS: Vineland Adaptive Behaviour Scale, ADHD: Attention Deficit/Hyperactivity Disorder, PPVT: Peabody Picture Vocabulary Task.

Overall, PRS-ASD only had a significant effect on 13 phenotypes. In brief, PRS-ASD influenced one core ASD feature (increased the severity of SCQ); increased non-verbal and full-scale IQ (cognitive ability); decreased the risk for intellectual disability (adaptive functioning); lowered the age of five developmental milestones (developmental features); decreased the risk for separation anxiety (co-occurring disorder) and word delay (language ability); increased the risk for sibling diagnosis of ASD (family history); and reduced the risk for nonfebrile seizures (neurological disorder).

In comparison, the multi-PRS dimensions had a significant effect on a substantial number of developmental phenotypes in ASD (Figure 5, Figure S8). Moreover, the multi-PRS dimensions that conferred significant genetic risk for ASD (Figure 5) had unique effects on core ASD and developmental features (Figure 6). Interestingly, we found a striking “mirroring” pattern in the effects of PC1 and PC2 on the risk for core ASD features, developmental features, and risk for co-occurring disorders. PC1, which mostly captured the PRS variability for negative symptom outcomes (Figure 4a: depressive symptoms, MDD, neuroticism, and tiredness), significantly increased the severity of core ASD features, the age of developmental milestones, and increased the risk for word delay and familial ASD. In contrast, PC2 – which mostly captured the PRS variability for high cognitive ability (Figure 4a: college and educational attainment, intelligence) – significantly reduced the severity of core ASD features, increased cognitive ability, reduced the age of developmental milestones, and decreased the risk for co-occurring disorders and word delay.

## DISCUSSION

This findings from this study suggest that the paucity of genome-wide significant hits in ASD may be partially driven by the complex and heterogenous genetic architecture of the disorder.

We applied PCA to 11 ASD-related PRSs to construct PRSs that captured the additive, inherited, and non-specific genetic risk for ASD in unique and orthogonal dimensions (referred to as “multi-PRS” dimensions).

First, we found that the multi-PRS dimensions can capture a similar proportion of genetic risk for ASD (0.501%), compared to PRS-ASD itself (0.619%). This suggests that ASD-related PRSs may be used as a proxy for PRS-ASD to capture a similar proportion of genetic risk for the disorder.

Second, we found that the multi-PRS approach can capture unique differences in developmental phenotypes among cases with ASD that PRS-ASD cannot. While multi-PRS dimensions significantly increased the risk for ASD, there was a striking “mirroring” effect between the PC1 and PC2 multi-PRS dimensions on core ASD features, developmental outcomes, and risk for co-occurring disorders. PC1 – which mostly captures the polygenic variability for negative symptom outcomes – increased the risk for ASD and increased the severity of core ASD features, delayed developmental milestones, and risk for co-occurring disorders Conversely, PC2 – which mostly captures the polygenic variability for higher cognitive ability – also increased the risk for ASD, yet in contrast, reduced the severity of core ASD features, was associated with earlier developmental milestones, and reduced the risk for co-occurring disorders. In other words, our findings highlight the heterogeneity underlying the genetic risk for ASD. While ASD-related PRSs substantially increase the risk for ASD, they have “opposing” effects on core and peripheral ASD phenotypes. This, in turn, could partially explain the low yield of genome-wide significant hits in ASD. We posit that this “mirroring” phenomenon may dilute the association signal of risk loci in ASD, and could partially explain the diversity observed among individuals with the disorder.

The findings from this study may also reflect the role of a general psychopathology factor (or “P-factor”) on ASD, which captures the variance across psychiatric symptoms in a shared dimension^43–45^. Our results indicate that multi-PRS dimensions can capture a significant, albeit small, proportion of the inherited genetic liability for ASD. Our proposed multi-PRS approach may represent a genetic “P-factor” that captures the overall liability for a diagnosis of a mental disorder^46, 47^. Several studies have aimed to identify a genomic P-factor that captures the general liability for psychopathology. Selzam *et al*. proposed a “polygenic P-factor” by applying PCA to PRSs for eight psychopathology traits and investigating the loadings of each trait on the first PC^46^. They found that this genomic P-factor explained 20-43% of the SNP effects across the disorders. Krapohl *et al*. modelled the effect of multiple PRSs on three developmental outcomes and found that combining multiple PRSs in a model yields better phenotype prediction than single-score predictor models^48^.

These findings also align with the Research Domain Criteria (RDoC), which represents a framework for re-classifying mental disorders based on dimensional behaviours and neurobiological measures^49^. Rather than focusing on binary categories, the RDoC examines the underlying pathophysiology of basic traits along a continuum^50^. Indeed, our findings highlight the benefit of studying the genetic architecture of ASD and its developmental phenotypes via related PRSs. While both the multi-PRS PC1 and PC2 dimensions significantly increase the risk for ASD, they have distinct – and opposing – effects on phenotypic characteristics among cases with ASD. In other words, these findings could not have been elucidated through a solely unitary approach (i.e.: the effect of PRS-ASD on ASD diagnosis alone).

We propose three major use cases for this novel multi-PRS approach. First, constructing a PRS from related traits can serve as a proxy for polygenic risk in studies where the GWAS summary statistics overlap with the individual-level genetic data. Second, this approach can be used to generate a PRS for traits that lack sufficient GWAS statistical power. Finally, the multi-PRS dimensions may account for pleiotropy and heterogeneity in orthogonal dimensions. This overcomes the need for conventional multivariate regression models that include all ASD-related PRSs as predictors and are thus subject to overfitting and multicollinearity.

This study has some limitations. First, while this intrafamilial study allows for the comparison of polygenic risk among affected and unaffected family members with ASD, we expect undiagnosed parents and siblings of probands with ASD to have higher rates of ASD traits as compared to the general population^51^. To account for this, we did not include in the analysis those parents and siblings that also had a diagnosis of ASD. However, further comparisons of phenotypes between individuals with ASD and their relatives were not possible due to limited phenotypic data available for relatives. Second, this study aggregates PRSs across numerous genotyping and sequencing technologies. To ensure that the PRSs across modalities were comparable, we only included loci present across all technologies before constructing the PRSs. We also compared PRSs (Figure S1, S2, S3) and applied PCA across various sensitivity analysis groups (Figure S4, S5, S6) and we found that the correlation structure and the main dimensions of variance of polygenic risk were robust across technologies and cohorts.

Using a developmental deconstruction approach, this study contributes to the proposed role of inherited, additive, and non-ASD-specific genetic risk factors on ASD and its related phenotypes. Our proposed multi-PRS approach highlights the pleiotropy^52^ between ASD and its related traits, which increase the risk for ASD and uniquely influence developmental phenotypes among cases with ASD. While the use of PRS for clinical risk assessment at the individual level remains ill-advised^53^, this paper highlights the heterogeneous common genetic architecture of ASD that may hinder GWAS loci discovery.

## STATEMENTS AND DECLARATIONS

### Ethics approval

This study was performed in line with the principles of the Declaration of Helsinki.

## Data Availability

All data produced in the present study are available upon reasonable request to the authors.

## TABLES AND FIGURES

