## Supplemental Files for "Exploring the common genetic architecture of autism spectrum disorder using a novel multi-polygenic risk score approach"

### Supplemental Figures

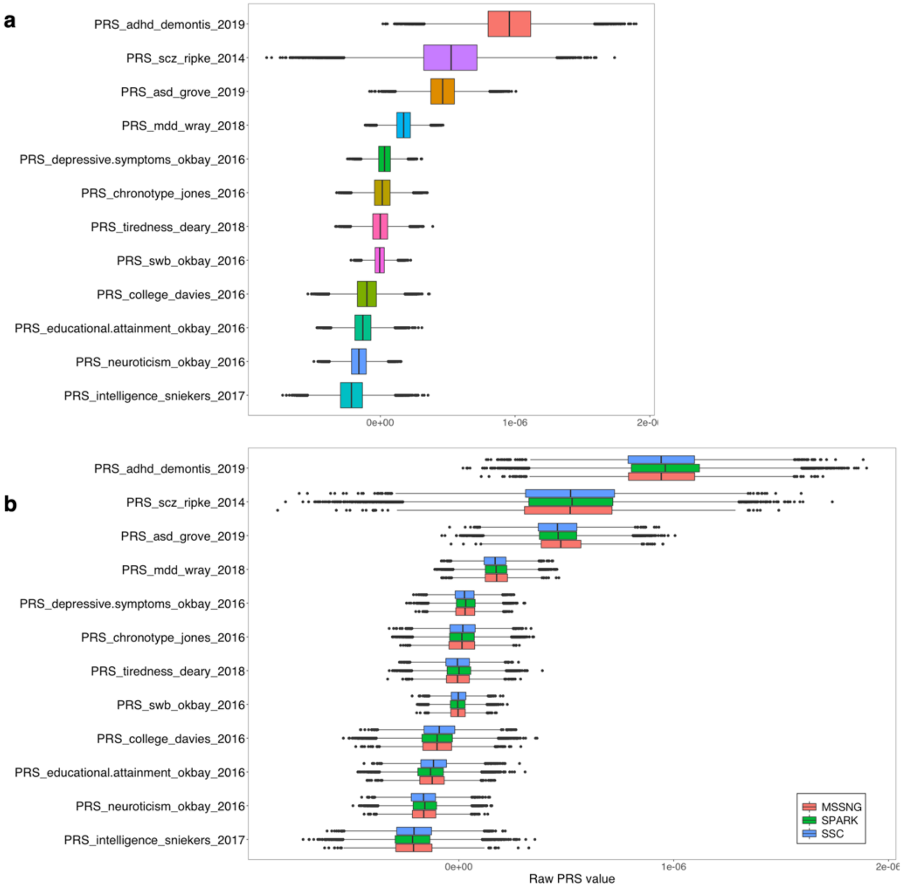


**Figure S1**. **Box plot highlighting the distribution of raw PRSs for all traits included in the study.** The raw PRS represents the PRS before accounting for the subtle differences in population structure among the samples (*see* Methods). a) The distribution of PRS for all traits included in the study. b) PRS distribution is not preferentially driven by a cohort. The distribution of PRS across the three (SSC, SPARK, MSSNG) ASD cohorts is comparable.

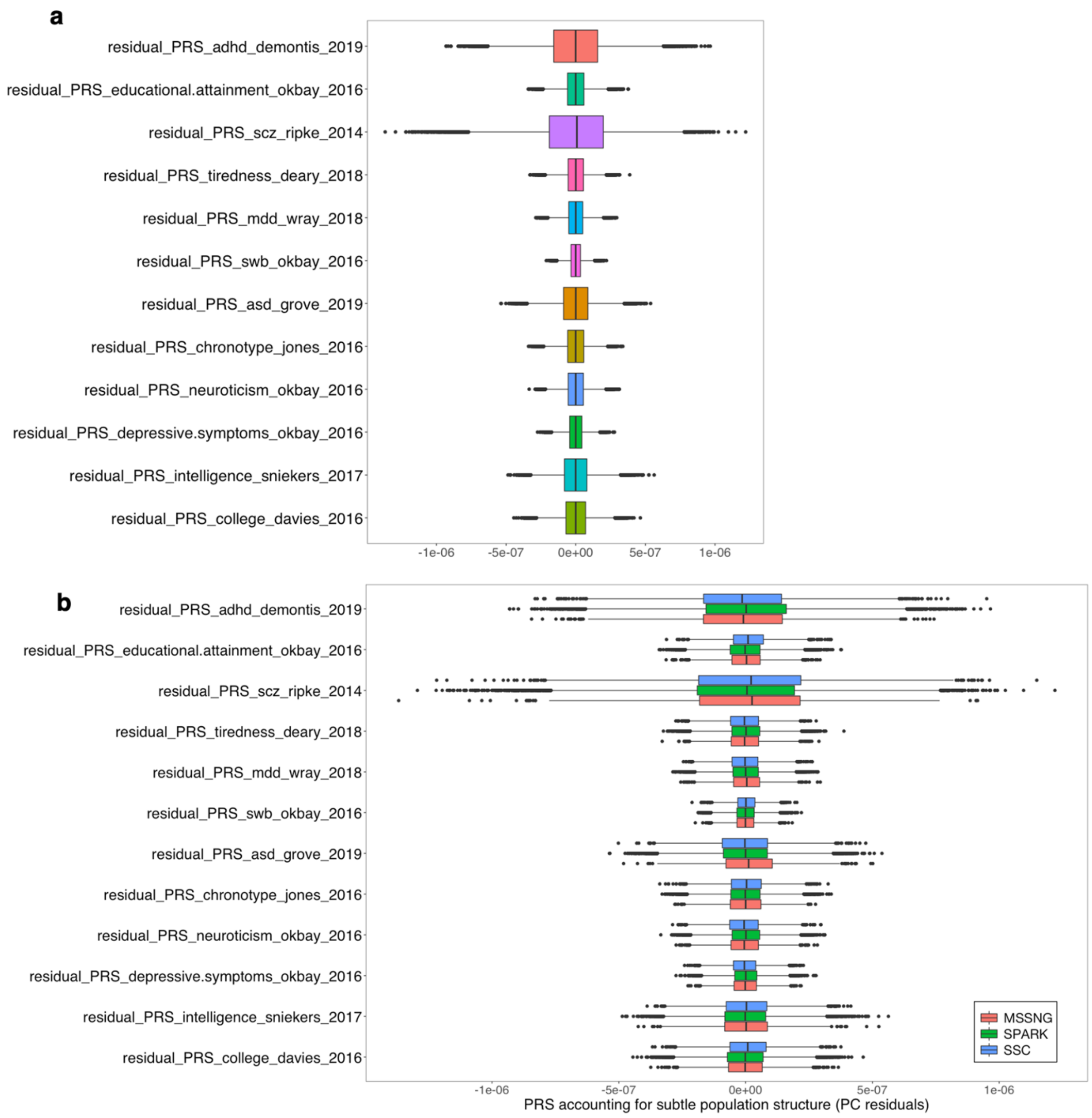


**Figure S2**. **Box plot highlighting the distribution of the PRS across all traits after accounting for ancestry.** Given the ancestry of the GWAS summary statistics included in the study, we restricted our analyses to European samples. To account for subtle differences in population structure, we regressed each raw PRS against the top 10 ancestry PCs of the European samples and extracted the residuals from these models. The residuals represented a PRS that removed the effects driven by ancestry. a) Distribution of the residuals (ancestry-accounted PRS) across all traits. b) Distribution of residuals across all traits for each cohort. There is no difference in distribution across the distinct ASD cohorts.

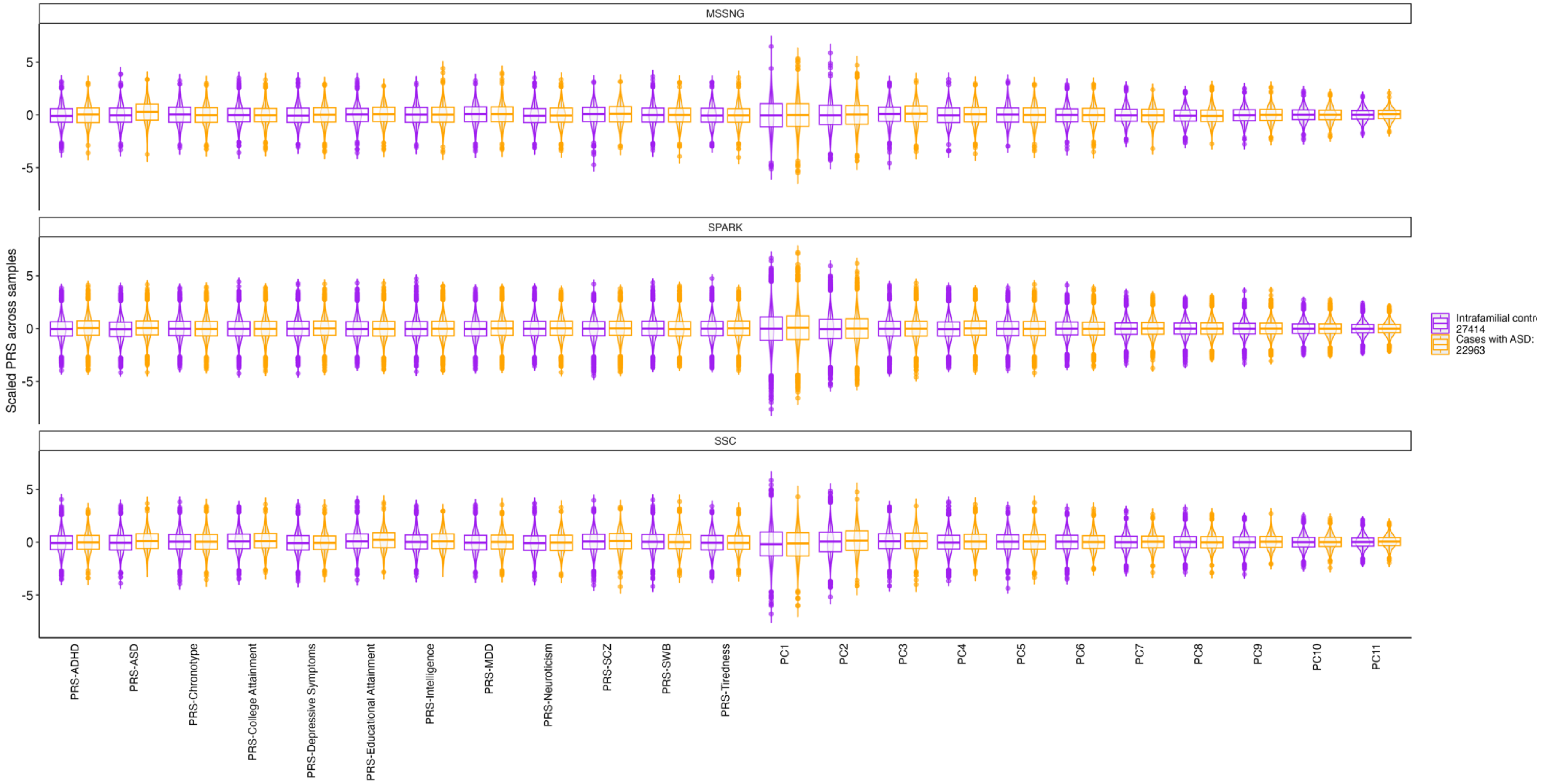


**Figure S3. Distribution of scaled PRSs and multi-PRS dimensions**. Cases with ASD (purple) and intrafamilial controls (orange) across the three different cohorts included in the study.

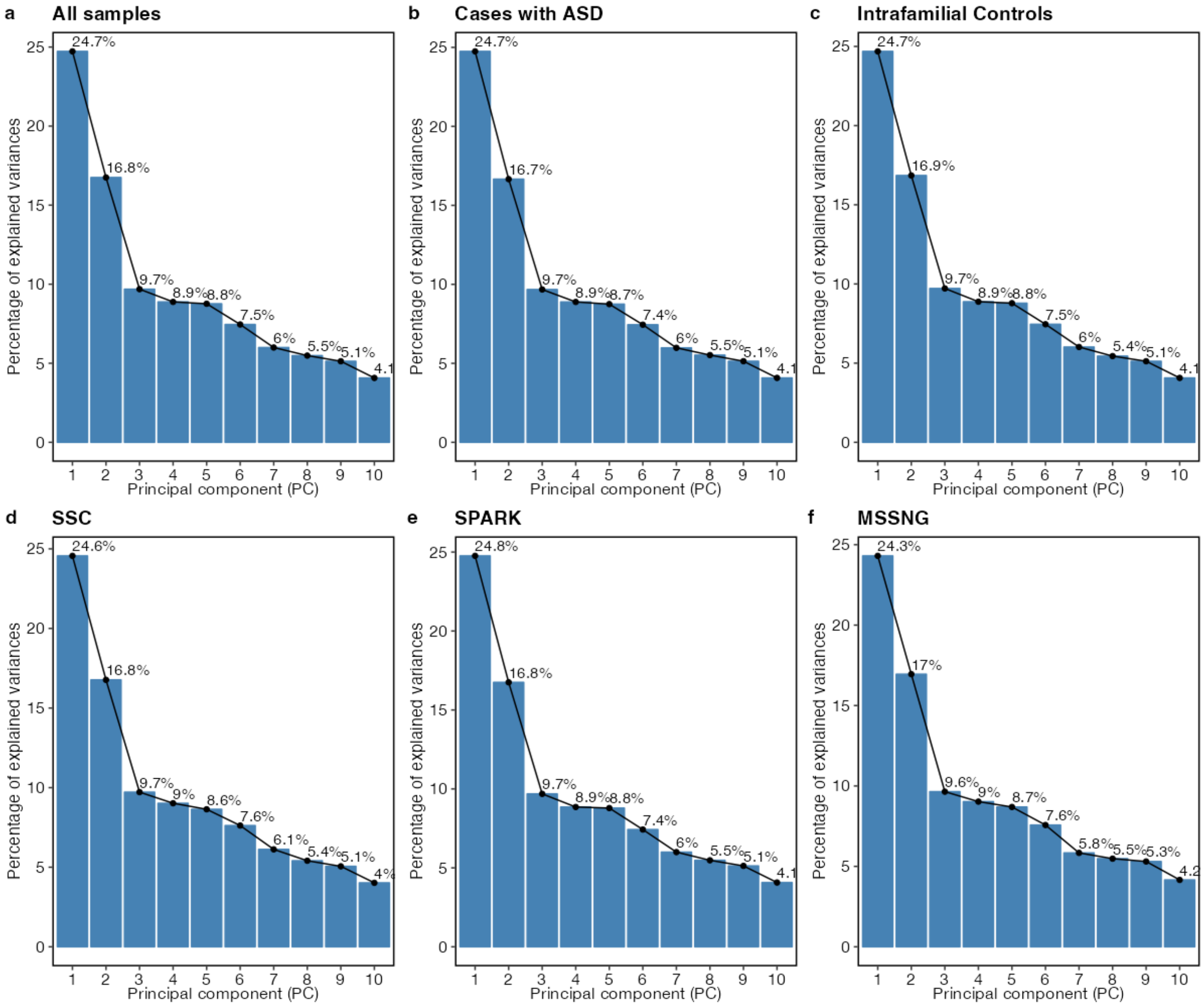


**Figure S4**. **PC variability across sensitivity analysis groups.** Compared to the main analyses presented in the manuscript (**a**), there is no discernable difference in the proportion of variability captured by each multi-PRS dimension (PC) across different analysis groups.

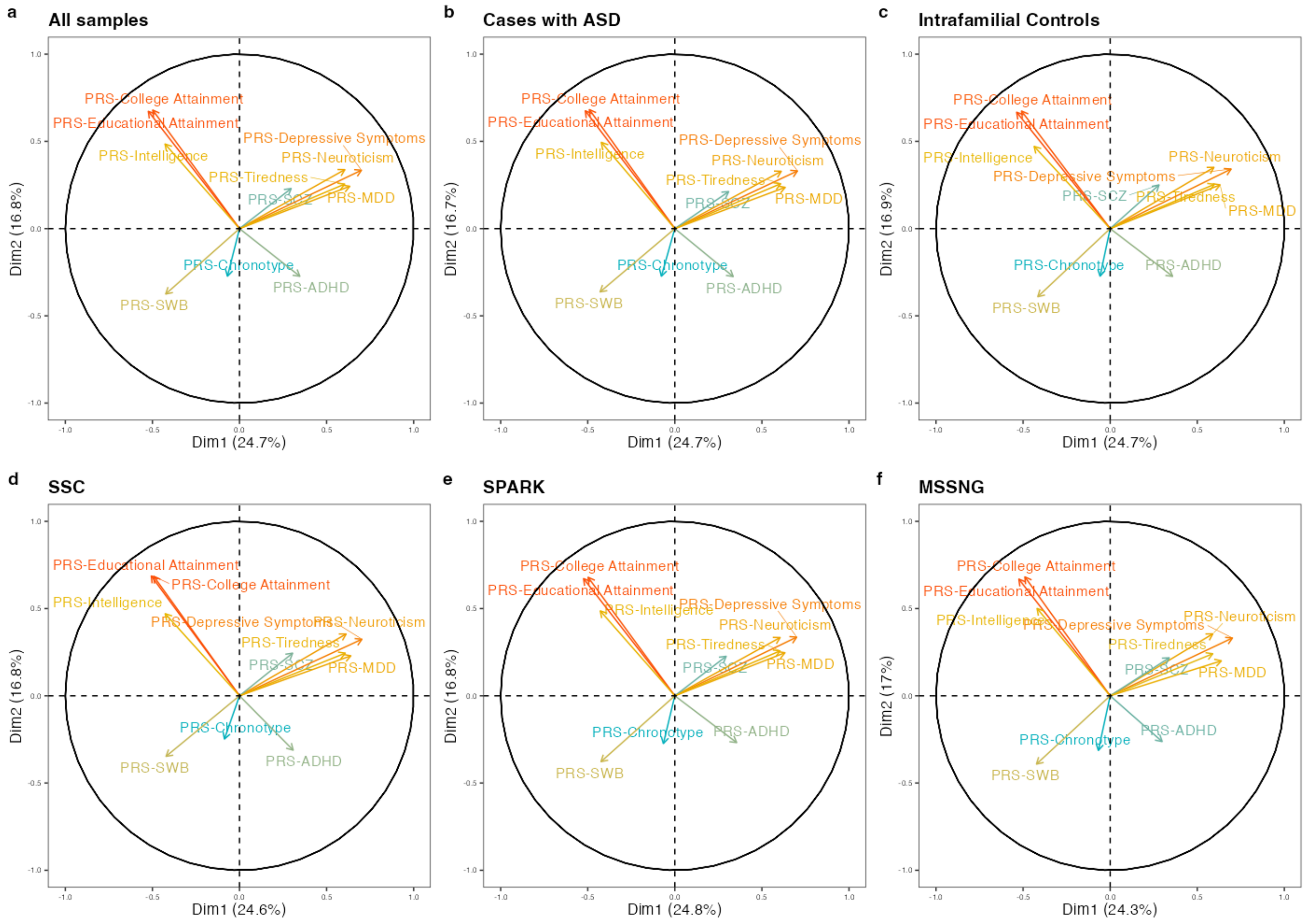


**Figure S5**. **PC1 versus PC2 relationship of ASD-related PRSs across sensitivity analysis groups.** Compared to the main analyses presented in the manuscript (**a**), there is no discernable difference in the relationship between ASD-related PRSs across different analysis groups. This sensitivity analysis suggests that the relationship between traits reported in the manuscript is not preferentially driven by either case-control status or cohort.

**a** **All samples**

| | PC1 | PC2 | PC3 | PC4 | PC5 | PC6 | PC7 | PC8 | PC9 | PC10 | PC11 |
|---|---|---|---|---|---|---|---|---|---|---|---|
| PRS-ADHD | 0.12 | 0.07 | 0.00 | 0.65 | 0.01 | 0.01 | 0.04 | 0.06 | 0.02 | 0.01 | 0.00 |
| PRS-Chronotype | 0.00 | 0.07 | 0.03 | 0.02 | 0.86 | 0.00 | 0.01 | 0.00 | 0.00 | 0.00 | 0.00 |
| PRS-Depressive Symptoms | 0.49 | 0.11 | 0.01 | 0.00 | 0.01 | 0.07 | 0.04 | 0.00 | 0.01 | 0.26 | 0.00 |
| PRS-College Attainment | 0.25 | 0.46 | 0.01 | 0.02 | 0.02 | 0.00 | 0.00 | 0.08 | 0.01 | 0.00 | 0.16 |
| PRS-Educational Attainment | 0.27 | 0.45 | 0.03 | 0.00 | 0.01 | 0.01 | 0.00 | 0.04 | 0.01 | 0.00 | 0.17 |
| PRS-Intelligence | 0.18 | 0.24 | 0.15 | 0.08 | 0.01 | 0.00 | 0.01 | 0.27 | 0.06 | 0.00 | 0.00 |
| PRS-MDD | 0.40 | 0.06 | 0.04 | 0.12 | 0.01 | 0.00 | 0.18 | 0.04 | 0.04 | 0.10 | 0.00 |
| PRS-Neuroticism | 0.37 | 0.11 | 0.08 | 0.05 | 0.02 | 0.00 | 0.02 | 0.07 | 0.25 | 0.03 | 0.00 |
| PRS-SCZ | 0.09 | 0.05 | 0.67 | 0.00 | 0.02 | 0.00 | 0.04 | 0.03 | 0.09 | 0.00 | 0.00 |
| PRS-SWB | 0.18 | 0.14 | 0.00 | 0.02 | 0.00 | 0.58 | 0.04 | 0.00 | 0.04 | 0.00 | 0.00 |
| PRS-Tiredness | 0.36 | 0.07 | 0.05 | 0.00 | 0.00 | 0.14 | 0.29 | 0.00 | 0.04 | 0.04 | 0.00 |

**b** **Cases with ASD**

| | PC1 | PC2 | PC3 | PC4 | PC5 | PC6 | PC7 | PC8 | PC9 | PC10 | PC11 |
|---|---|---|---|---|---|---|---|---|---|---|---|
| PRS-ADHD | 0.11 | 0.08 | 0.00 | 0.66 | 0.00 | 0.02 | 0.04 | 0.06 | 0.01 | 0.01 | 0.00 |
| PRS-Chronotype | 0.01 | 0.07 | 0.03 | 0.01 | 0.86 | 0.01 | 0.00 | 0.00 | 0.00 | 0.00 | 0.00 |
| PRS-Depressive Symptoms | 0.50 | 0.11 | 0.01 | 0.00 | 0.01 | 0.07 | 0.04 | 0.00 | 0.01 | 0.26 | 0.00 |
| PRS-College Attainment | 0.24 | 0.47 | 0.02 | 0.02 | 0.01 | 0.00 | 0.00 | 0.07 | 0.01 | 0.00 | 0.16 |
| PRS-Educational Attainment | 0.26 | 0.46 | 0.03 | 0.00 | 0.00 | 0.01 | 0.00 | 0.05 | 0.01 | 0.00 | 0.18 |
| PRS-Intelligence | 0.18 | 0.24 | 0.13 | 0.09 | 0.01 | 0.00 | 0.01 | 0.27 | 0.06 | 0.00 | 0.00 |
| PRS-MDD | 0.40 | 0.06 | 0.05 | 0.11 | 0.01 | 0.00 | 0.19 | 0.04 | 0.04 | 0.10 | 0.00 |
| PRS-Neuroticism | 0.37 | 0.11 | 0.08 | 0.04 | 0.02 | 0.00 | 0.02 | 0.08 | 0.25 | 0.03 | 0.00 |
| PRS-SCZ | 0.10 | 0.04 | 0.65 | 0.01 | 0.02 | 0.00 | 0.04 | 0.03 | 0.09 | 0.00 | 0.00 |
| PRS-SWB | 0.18 | 0.13 | 0.00 | 0.03 | 0.01 | 0.56 | 0.03 | 0.00 | 0.04 | 0.00 | 0.00 |
| PRS-Tiredness | 0.37 | 0.07 | 0.05 | 0.00 | 0.00 | 0.15 | 0.28 | 0.00 | 0.04 | 0.04 | 0.00 |

**c** **Intrafamilial Controls**

| | PC1 | PC2 | PC3 | PC4 | PC5 | PC6 | PC7 | PC8 | PC9 | PC10 | PC11 |
|---|---|---|---|---|---|---|---|---|---|---|---|
| PRS-ADHD | 0.13 | 0.07 | 0.00 | 0.59 | 0.05 | 0.01 | 0.04 | 0.07 | 0.02 | 0.01 | 0.00 |
| PRS-Chronotype | 0.00 | 0.07 | 0.02 | 0.10 | 0.79 | 0.00 | 0.01 | 0.00 | 0.00 | 0.00 | 0.00 |
| PRS-Depressive Symptoms | 0.48 | 0.12 | 0.02 | 0.01 | 0.01 | 0.06 | 0.04 | 0.00 | 0.02 | 0.25 | 0.00 |
| PRS-College Attainment | 0.26 | 0.45 | 0.01 | 0.01 | 0.02 | 0.01 | 0.00 | 0.08 | 0.01 | 0.00 | 0.15 |
| PRS-Educational Attainment | 0.29 | 0.45 | 0.02 | 0.00 | 0.01 | 0.01 | 0.00 | 0.04 | 0.01 | 0.00 | 0.17 |
| PRS-Intelligence | 0.19 | 0.22 | 0.17 | 0.06 | 0.02 | 0.00 | 0.00 | 0.26 | 0.06 | 0.00 | 0.00 |
| PRS-MDD | 0.40 | 0.06 | 0.03 | 0.11 | 0.04 | 0.00 | 0.16 | 0.04 | 0.04 | 0.11 | 0.00 |
| PRS-Neuroticism | 0.36 | 0.12 | 0.07 | 0.07 | 0.00 | 0.00 | 0.02 | 0.07 | 0.24 | 0.03 | 0.00 |
| PRS-SCZ | 0.08 | 0.06 | 0.67 | 0.00 | 0.01 | 0.01 | 0.03 | 0.03 | 0.09 | 0.01 | 0.00 |
| PRS-SWB | 0.17 | 0.15 | 0.00 | 0.01 | 0.00 | 0.58 | 0.04 | 0.00 | 0.04 | 0.00 | 0.00 |
| PRS-Tiredness | 0.36 | 0.06 | 0.05 | 0.01 | 0.00 | 0.14 | 0.31 | 0.00 | 0.03 | 0.04 | 0.00 |

**d** **SSC**

| | PC1 | PC2 | PC3 | PC4 | PC5 | PC6 | PC7 | PC8 | PC9 | PC10 | PC11 |
|---|---|---|---|---|---|---|---|---|---|---|---|
| PRS-ADHD | 0.10 | 0.10 | 0.01 | 0.63 | 0.04 | 0.00 | 0.01 | 0.09 | 0.01 | 0.01 | 0.00 |
| PRS-Chronotype | 0.01 | 0.06 | 0.14 | 0.08 | 0.69 | 0.02 | 0.00 | 0.00 | 0.00 | 0.00 | 0.00 |
| PRS-Depressive Symptoms | 0.49 | 0.11 | 0.01 | 0.00 | 0.00 | 0.09 | 0.02 | 0.01 | 0.00 | 0.25 | 0.00 |
| PRS-College Attainment | 0.24 | 0.47 | 0.02 | 0.02 | 0.00 | 0.01 | 0.00 | 0.07 | 0.01 | 0.00 | 0.16 |
| PRS-Educational Attainment | 0.26 | 0.47 | 0.03 | 0.00 | 0.00 | 0.01 | 0.00 | 0.03 | 0.02 | 0.00 | 0.17 |
| PRS-Intelligence | 0.18 | 0.22 | 0.10 | 0.09 | 0.08 | 0.01 | 0.04 | 0.21 | 0.07 | 0.00 | 0.00 |
| PRS-MDD | 0.41 | 0.05 | 0.06 | 0.11 | 0.01 | 0.00 | 0.11 | 0.09 | 0.08 | 0.08 | 0.00 |
| PRS-Neuroticism | 0.37 | 0.13 | 0.07 | 0.03 | 0.02 | 0.00 | 0.07 | 0.06 | 0.21 | 0.04 | 0.00 |
| PRS-SCZ | 0.09 | 0.06 | 0.60 | 0.00 | 0.07 | 0.00 | 0.05 | 0.02 | 0.10 | 0.00 | 0.00 |
| PRS-SWB | 0.18 | 0.12 | 0.00 | 0.03 | 0.03 | 0.57 | 0.02 | 0.01 | 0.03 | 0.01 | 0.00 |
| PRS-Tiredness | 0.37 | 0.06 | 0.04 | 0.00 | 0.00 | 0.12 | 0.33 | 0.01 | 0.03 | 0.04 | 0.00 |

**e** **SPARK**

| | PC1 | PC2 | PC3 | PC4 | PC5 | PC6 | PC7 | PC8 | PC9 | PC10 | PC11 |
|---|---|---|---|---|---|---|---|---|---|---|---|
| PRS-ADHD | 0.13 | 0.07 | 0.00 | 0.64 | 0.01 | 0.01 | 0.05 | 0.06 | 0.02 | 0.01 | 0.00 |
| PRS-Chronotype | 0.00 | 0.07 | 0.01 | 0.02 | 0.87 | 0.00 | 0.01 | 0.00 | 0.00 | 0.00 | 0.00 |
| PRS-Depressive Symptoms | 0.49 | 0.11 | 0.01 | 0.00 | 0.01 | 0.06 | 0.04 | 0.00 | 0.02 | 0.26 | 0.00 |
| PRS-College Attainment | 0.25 | 0.46 | 0.01 | 0.02 | 0.02 | 0.00 | 0.00 | 0.08 | 0.01 | 0.00 | 0.16 |
| PRS-Educational Attainment | 0.27 | 0.45 | 0.03 | 0.00 | 0.01 | 0.01 | 0.00 | 0.04 | 0.01 | 0.00 | 0.17 |
| PRS-Intelligence | 0.18 | 0.24 | 0.16 | 0.08 | 0.01 | 0.00 | 0.00 | 0.27 | 0.06 | 0.00 | 0.00 |
| PRS-MDD | 0.40 | 0.06 | 0.04 | 0.12 | 0.02 | 0.00 | 0.19 | 0.04 | 0.04 | 0.11 | 0.00 |
| PRS-Neuroticism | 0.37 | 0.11 | 0.08 | 0.06 | 0.01 | 0.00 | 0.02 | 0.08 | 0.25 | 0.03 | 0.00 |
| PRS-SCZ | 0.09 | 0.05 | 0.68 | 0.00 | 0.01 | 0.00 | 0.03 | 0.04 | 0.09 | 0.01 | 0.00 |
| PRS-SWB | 0.18 | 0.14 | 0.00 | 0.03 | 0.00 | 0.57 | 0.03 | 0.00 | 0.04 | 0.00 | 0.00 |
| PRS-Tiredness | 0.37 | 0.07 | 0.05 | 0.00 | 0.00 | 0.14 | 0.29 | 0.00 | 0.04 | 0.04 | 0.00 |

**f** **MSSNG**

| | PC1 | PC2 | PC3 | PC4 | PC5 | PC6 | PC7 | PC8 | PC9 | PC10 | PC11 |
|---|---|---|---|---|---|---|---|---|---|---|---|
| PRS-ADHD | 0.09 | 0.07 | 0.03 | 0.69 | 0.00 | 0.00 | 0.05 | 0.02 | 0.04 | 0.01 | 0.00 |
| PRS-Chronotype | 0.00 | 0.10 | 0.02 | 0.00 | 0.87 | 0.00 | 0.01 | 0.00 | 0.01 | 0.00 | 0.00 |
| PRS-Depressive Symptoms | 0.49 | 0.11 | 0.01 | 0.00 | 0.01 | 0.05 | 0.05 | 0.00 | 0.00 | 0.27 | 0.00 |
| PRS-College Attainment | 0.24 | 0.46 | 0.01 | 0.03 | 0.01 | 0.01 | 0.00 | 0.07 | 0.00 | 0.00 | 0.16 |
| PRS-Educational Attainment | 0.27 | 0.45 | 0.02 | 0.01 | 0.01 | 0.02 | 0.00 | 0.05 | 0.00 | 0.00 | 0.17 |
| PRS-Intelligence | 0.18 | 0.25 | 0.17 | 0.04 | 0.00 | 0.01 | 0.00 | 0.35 | 0.00 | 0.00 | 0.00 |
| PRS-MDD | 0.41 | 0.04 | 0.04 | 0.15 | 0.00 | 0.00 | 0.19 | 0.01 | 0.08 | 0.09 | 0.00 |
| PRS-Neuroticism | 0.35 | 0.13 | 0.07 | 0.05 | 0.04 | 0.01 | 0.01 | 0.00 | 0.30 | 0.04 | 0.00 |
| PRS-SCZ | 0.12 | 0.05 | 0.63 | 0.00 | 0.01 | 0.01 | 0.06 | 0.08 | 0.05 | 0.00 | 0.00 |
| PRS-SWB | 0.18 | 0.15 | 0.00 | 0.01 | 0.00 | 0.54 | 0.06 | 0.01 | 0.05 | 0.00 | 0.00 |
| PRS-Tiredness | 0.35 | 0.06 | 0.07 | 0.01 | 0.00 | 0.19 | 0.22 | 0.00 | 0.06 | 0.04 | 0.00 |

**Figure S6**. **ASD-related PRS contribution to PCs across sensitivity analysis groups.** Compared to the main analyses presented in the manuscript (**a**), there is no discernable difference in the proportion of PRS for ASD-related traits, captured by the multi-PRS dimensions, across different analysis groups. These sensitivity analyses suggest that the results from the PCA are not preferentially driven by different sample sizes, case-control status, or cohort.

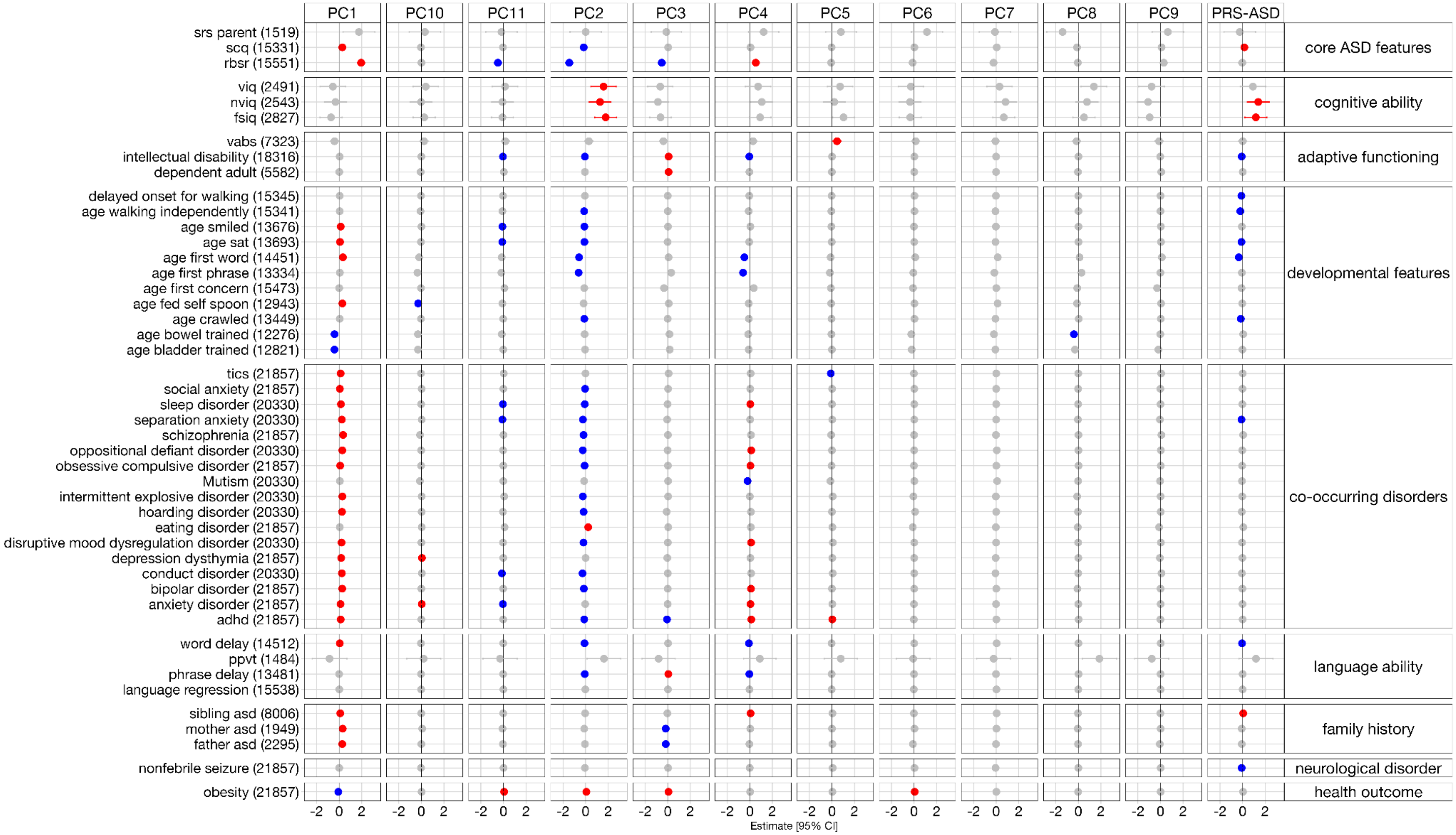


**Figure S7**. The effect of PRS-ASD, all ASD-related PRSs, and all multi-PRS dimensions on developmental phenotypes among cases with ASD. The effect of PRS-ASD and its related traits on each clinical outcome were modelled independently. All multi-PRS dimensions were included as predictors in the same model for each clinical outcome. The effect of each predictor on the clinical outcome is also detailed in Supplementary Table

5. Red and blue dots represent a significant positive and negative effect of the PRS predictor variable on the clinical outcome, respectively. All *P* values were adjusted for FDR correction. The brackets denote the number of cases with ASD that were included in the model.
